# *ApoE* e4e4 genotype and mortality with COVID-19 in UK Biobank

**DOI:** 10.1101/2020.06.19.20134908

**Authors:** Chia-Ling Kuo, Luke C Pilling, Janice L Atkins, Jane AH Masoli, João Delgado, George A Kuchel, David Melzer

## Abstract

We previously reported that the *ApoE* e4e4 genotype was associated with COVID-19 test positivity (OR=2.31, 95% CI: 1.65 to 3.24, p=1.19×10^−6^) in the UK Biobank (UKB) cohort, during the epidemic peak in England, from March 16 to April 26, 2020. With more COVID-19 test results (March 16 to May 31, 2020) and mortality data (to April 26, 2020) linked to UKB, we re-evaluated the *ApoE* e4 allele association with COVID-19 test positivity, and with all-cause mortality following test-confirmed COVID-19. Logistic regression models compared *ApoE* e4e4 participants (or e3e4s) to e3e3s with adjustment for sex; age on April 26th or age at death; baseline UKB assessment center in England (accounting for geographical differences in viral exposures); genotyping array type; and the top five genetic principal components (accounting for possible population admixture). *ApoE* e4e4 genotype was associated with increased risks of test positivity (OR=2.24, 95% CI: 1.72 to 2.93, p=3.24×10^−9^) and of mortality with test-confirmed COVID-19 (OR=4.29, 95% CI: 2.38 to 7.72, p=1.22×10^−6^), compared to e3e3s. Independent replications are needed to confirm our findings and mechanistic work is needed to understand how *ApoE* e4e4 results in the marked increase in vulnerability, especially for COVID-19 mortality. These findings also demonstrate that risks for COVID-19 mortality are not simply related to advanced chronological age or the comorbidities commonly seen in aging.

## Letter to Editor

We previously reported that the *ApoE* e4e4 genotype was associated with COVID-19 test positivity (OR=2.31, 95% CI: 1.65 to 3.24, p=1.19×10^−6^) [1] in the UK Biobank (UKB) cohort, during the epidemic peak in England, from March 16 to April 26, 2020 [2]. With more COVID-19 test results (March 16 to May 31, 2020) and mortality data (to April 26, 2020) linked to UKB, we re-evaluated the *ApoE* e4 allele association with COVID-19 test positivity, and with all-cause mortality following test-confirmed COVID-19.

We restricted analyses to European-ancestry participants [3] (n=451,367, 90% of sample) attending baseline assessment centers in England (n=398,073) and excluded participants who died before the pandemic (set at February 1, 2020, n=22,384). Single nucleotide polymorphism data for rs429358 and rs7412 were used to determine *ApoE* genotypes. Our outcomes of interest were: a) COVID-19 test positive versus the rest of the sample meeting inclusion criteria (i.e., including untested samples and tested negative), and b) tested positive and died versus the rest of the sample as above, but with additional exclusion of test positive participants who survived. Logistic regression models compared *ApoE* e4e4 participants (or e3e4s) to e3e3s with adjustment for sex; age on April 26th or age at death; baseline UKB assessment center in England (accounting for geographical differences in viral exposures); genotyping array type; and the top five genetic principal components (accounting for possible population admixture).

The mean attained age was 68.2 years (SD=8.0) with 174,667 females (55%). Of 219,747 e3e3 participants, 663 participants tested positive (302 per 100,000), of whom, 79 later died. Similarly, of 8,767 e4e4 participants, 59 tested positive (673 per 100,000), of whom 13 later died (Table 1). In logistic models, *ApoE* e4e4 genotype was associated with increased risks of test positivity (OR=2.24, 95% CI: 1.72 to 2.93, p=3.24×10^−9^) and of mortality with test-confirmed COVID-19 (OR=4.29, 95% CI: 2.38 to 7.72, p=1.22×10^−6^), compared to e3e3s. For e3e4s versus e3e3s, these two associations were nominally statistically significant (at p<0.05), but with much smaller effect sizes. The e4e4 associations were similar after excluding 50,566 participants related to the 3^rd^-degree or closer for test positivity (e4e4 OR=2.30, 95% CI: 1.73 to 3.07, p=1.39×10^−8^) and for mortality with test-confirmed COVID-19 (e4e4 OR=4.53, 95% CI: 2.39 to 8.61, p=3.87×10^−6^). Additionally, the e4e4 association with either COVID-19 outcome was little changed after removing participants with diseases associated with *ApoE* e4 alleles [5] and COVID-19 severity [6], including dementia, hypertension, coronary artery disease (myocardial infarction or angina), or type 2 diabetes (Table 1), based on diagnoses recorded from baseline self-reports and hospital discharge records during follow-up to March 2017. *ApoE* e3e4s were modestly associated with test positivity overall, and the association tended to be less marked in disease-free samples (Table 1). In additional analyses, we tested associations with *ApoE* e2 alleles, which have been linked to beneficial health outcomes [5]. No associations were found between e2e3 and either of our COVID-19 outcomes (p>0.05, versus e3e3). Analyses for e2e2s associations were underpowered (n=2,427, 4 positives, and 1 positive death).

**Table 1.**
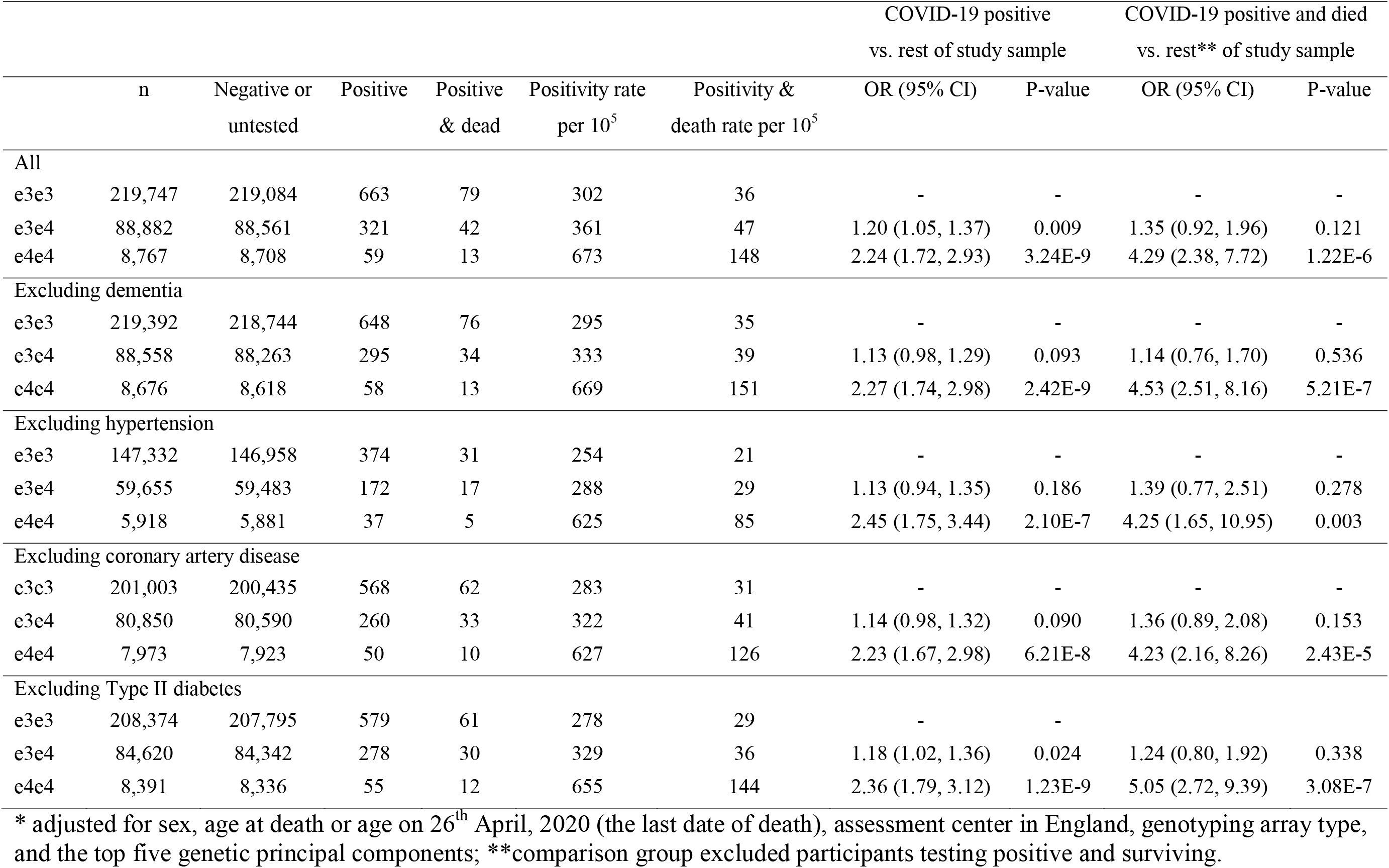
Risk of COVID-19 test positivity and mortality, comparing participants with *ApoE* e3e4 or e4e4 to e3e3 genotypes, in UK Biobank.

|  |  |  |  |  |  |  | COVID-19 positive<br>vs. rest of study sample |  | COVID-19 positive and died<br>vs. rest** of study sample |  |
| --- | --- | --- | --- | --- | --- | --- | --- | --- | --- | --- |
|  | n | Negative or<br>untested | Positive | Positive<br>& dead | Positivity rate<br>per 10<sup>5</sup> | Positivity &<br>death rate per 10<sup>5</sup> | OR (95% CI) | P-value | OR (95% CI) | P-value |
| All |  |  |  |  |  |  |  |  |  |  |
| e3e3 | 219,747 | 219,084 | 663 | 79 | 302 | 36 | - | - | - | - |
| e3e4 | 88,882 | 88,561 | 321 | 42 | 361 | 47 | 1.20 (1.05, 1.37) | 0.009 | 1.35 (0.92, 1.96) | 0.121 |
| e4e4 | 8,767 | 8,708 | 59 | 13 | 673 | 148 | 2.24 (1.72, 2.93) | 3.24E-9 | 4.29 (2.38, 7.72) | 1.22E-6 |
| Excluding dementia |  |  |  |  |  |  |  |  |  |  |
| e3e3 | 219,392 | 218,744 | 648 | 76 | 295 | 35 | - | - | - | - |
| e3e4 | 88,558 | 88,263 | 295 | 34 | 333 | 39 | 1.13 (0.98, 1.29) | 0.093 | 1.14 (0.76, 1.70) | 0.536 |
| e4e4 | 8,676 | 8,618 | 58 | 13 | 669 | 151 | 2.27 (1.74, 2.98) | 2.42E-9 | 4.53 (2.51, 8.16) | 5.21E-7 |
| Excluding hypertension |  |  |  |  |  |  |  |  |  |  |
| e3e3 | 147,332 | 146,958 | 374 | 31 | 254 | 21 | - | - | - | - |
| e3e4 | 59,655 | 59,483 | 172 | 17 | 288 | 29 | 1.13 (0.94, 1.35) | 0.186 | 1.39 (0.77, 2.51) | 0.278 |
| e4e4 | 5,918 | 5,881 | 37 | 5 | 625 | 85 | 2.45 (1.75, 3.44) | 2.10E-7 | 4.25 (1.65, 10.95) | 0.003 |
| Excluding coronary artery disease |  |  |  |  |  |  |  |  |  |  |
| e3e3 | 201,003 | 200,435 | 568 | 62 | 283 | 31 | - | - | - | - |
| e3e4 | 80,850 | 80,590 | 260 | 33 | 322 | 41 | 1.14 (0.98, 1.32) | 0.090 | 1.36 (0.89, 2.08) | 0.153 |
| e4e4 | 7,973 | 7,923 | 50 | 10 | 627 | 126 | 2.23 (1.67, 2.98) | 6.21E-8 | 4.23 (2.16, 8.26) | 2.43E-5 |
| Excluding Type II diabetes |  |  |  |  |  |  |  |  |  |  |
| e3e3 | 208,374 | 207,795 | 579 | 61 | 278 | 29 | - | - | - | - |
| e3e4 | 84,620 | 84,342 | 278 | 30 | 329 | 36 | 1.18 (1.02, 1.36) | 0.024 | 1.24 (0.80, 1.92) | 0.338 |
| e4e4 | 8,391 | 8,336 | 55 | 12 | 655 | 144 | 2.36 (1.79, 3.12) | 1.23E-9 | 5.05 (2.72, 9.39) | 3.08E-7 |

\* adjusted for sex, age at death or age on 26<sup>th</sup> April, 2020 (the last date of death), assessment center in England, genotyping array type, and the top five genetic principal components; \*\*comparison group excluded participants testing positive and surviving.

The results presented imply a recessive effect of the *ApoE* e4 allele. Only modest associations were present between the much more common e3e4 genotype and COVID-19 outcomes, similar to results for rs429358 (which separates 0, 1, and 2 copies of e4 alleles, OR=1.3, p=0.0026) reported for severe COVID-19 with respiratory failure in a recent additive effect genome-wide analysis [7]. *ApoE* e4e4 associations with test positivity and mortality were little affected by excluding dementia and other *ApoE* e4 associated diagnoses reported before March 2017: future work should include recent pre-existing diagnoses. More data are needed on *ApoE* and COVID-19 associations in other ancestry groups, as numbers of UK Biobank participants of such groups are unfortunately too small for this analysis.

In conclusion, *ApoE* e4e4 genotype is associated with COVID-19 test positivity at genome-wide significance (i.e., p<5×10^−8^) in UK Biobank, using data covering a longer period than previously reported. Similarly, the e4e4 genotype was associated with a four-fold increase in mortality after testing positive for COVID-19, in UK Biobank. Independent replications are needed to confirm our findings and mechanistic work is needed to understand how *ApoE* e4e4 results in the marked increase in vulnerability, especially for COVID-19 mortality. These findings also demonstrate that risks for COVID-19 mortality are not simply related to advanced chronological age or the comorbidities commonly seen in aging.

## Data Availability

This research was conducted using the UK Biobank resource, under the application 14631.

https://www.ukbiobank.ac.uk

## References

1. Kuo C-L, Pilling LC, Atkins JL, Masoli JAH, Delgado J, Kuchel GA, Melzer D. APOE e4 genotype predicts severe COVID-19 in the UK Biobank community cohort. Journals Gerontol Ser A [Internet]. 2020;. Available from: https://doi.org/10.1093/gerona/glaa131

2. Armstrong J, Rudkin JK, Allen N, Crook DW, Wilson D, Wyllie DH, O’Connell A-M. Dynamic linkage of COVID-19 test results between Public Health England’s Second Generation Surveillance System and UK Biobank. figshare. 2020;.

3. Pilling LC, Tamosauskaite J, Jones G, Wood AR, Jones L, Kuo CL, Kuchel GA, Ferrucci L, Melzer D. Common conditions associated with hereditary haemochromatosis genetic variants: Cohort study in UK Biobank. BMJ. 2019; 364.

4. World Health Organization. COVID-19 coding in ICD-10. 2020.

5. Kuo C-L, Pilling LC, Atkins JL, Kuchel GA, Melzer D. ApoE e2 and aging-related outcomes in 379,000 UK Biobank participants. Aging (Albany NY) [Internet]. 2020;. Available from: https://doi.org/10.18632/aging

6. Atkins JL, Masoli JAH, Delgado J, Pilling LC, Kuo C-LC, Kuchel G, Melzer D. PREEXISTING COMORBIDITIES PREDICTING SEVERE COVID-19 IN OLDER ADULTS IN THE UK BIOBANK COMMUNITY COHORT. medRxiv [Internet]. 2020;: 2020.05.06.20092700. Available from: http://medrxiv.org/content/early/2020/05/08/2020.05.06.20092700.abstract

7. Ellinghaus D, Degenhardt F, Bujanda L, Buti M, Albillos A, Invernizzi P, Fernández J, Prati D, Baselli G, Asselta R, Grimsrud MM, Milani C, Aziz F, et al. Genomewide Association Study of Severe Covid-19 with Respiratory Failure. N Engl J Med [Internet]. Massachusetts Medical Society; 2020;. Available from: https://doi.org/10.1056/NEJMoa2020283

